# Computational modeling of patient-specific healing and deformation outcomes following breast-conserving surgery based on MRI data

**DOI:** 10.1101/2025.08.06.25331892

**Authors:** Zachary Harbin, Carla Fisher, Sherry Voytik-Harbin, Adrian Buganza Tepole

## Abstract

**Purpose:** Breast-conserving surgery (BCS) is the standard of care for early-stage breast cancer, offering recurrence and survival rates comparable to mastectomy while preserving healthy breast tissue. However, surgical cavity healing post-BCS often leads to highly variable tissue remodeling, including scar tissue formation and contracture, leading to visible breast deformation or asymmetry. These outcomes significantly impact patient quality of life but are difficult to predict due to the complex interplay between biological healing processes and individual patient variability. To address this challenge, we extended our previously calibrated computational mechanobiological model of post-BCS healing by incorporating diagnostic imaging data to evaluate how patient-specific breast and tumor characteristics shape healing trajectories and breast deformation.

**Methods:** The model captured multiscale biological and biomechanical processes, including fibroblast activity, collagen remodeling, and nonlinear tissue mechanics, to simulate time-dependent tissue remodeling. Preoperative magnetic resonance imaging (MRI) scans provided patient-specific breast and tumor geometries and characteristics, which were integrated into finite element simulations of cavity healing. Simulation outputs were used to train Gaussian process surrogate models, enabling rapid, accurate prediction of healing dynamics and breast surface deformation across diverse patient profiles.

**Results:** These models revealed how factors including breast density, cavity volume, breast volume, and cavity depth influence post-surgical cavity contraction and measures of breast surface deformation.

**Conclusion:** This framework has the potential to provide a personalized, predictive tool for surgical planning and decision-making, enabling clinicians and patients to anticipate healing trajectories and cosmetic outcomes, with the goal of optimizing surgical results and enhancing patient quality of life.

## 1 Introduction

Breast cancer is among the most common cancers in women, with roughly 1 in 8 women diagnosed during their lifetime and over 367,000 new cases reported annually in the United States [1]. Increased awareness, routine screenings that enable early detection, and advancements in treatment have significantly improved survival rates, with localized breast cancer now having a 5-year survival rate exceeding 99% [1]. With these improved outcomes, clinical focus is shifting toward long-term considerations, including recurrence prevention and enhancement of quality of life.

Surgical treatments are associated with the lowest recurrence rates [2, 3] and are typically performed via breast-conserving surgery (BCS; otherwise known as lumpectomy) or mastectomy (complete removal of the breast). Choosing between these options is often complex and emotionally challenging, involving personal preferences, patient and cancer characteristics, and anticipated post-surgical outcomes. In recent years, BCS has emerged as the preferred standard of care for early-stage breast cancer, offering recurrence and survival rates comparable to mastectomy while carrying a lower risk of complications [4–8]. The primary goal of BCS is to completely remove the cancer while preserving as much healthy breast tissue and appearance as possible. However, the resulting surgical cavity initiates a complex healing process, resulting in variable levels of tissue contraction, scar tissue formation, and breast deformation (i.e., cosmetic defects, including dents, distortions, and asymmetries between breasts). Achieving a favorable cosmetic outcome is a critical component of surgical decision-making, as it is strongly associated with psychological recovery and longterm quality of life [9, 10]. Despite its importance, predicting oncologic, healing, and cosmetic outcomes remains challenging due to the complex nature of breast tissue repair and inter-patient variability. Factors such as breast and tumor geometry, tumor location, and breast density are known to influence healing, yet their contributions remain poorly understood. A deeper understanding of how these patient-specific characteristics affect BCS outcomes is needed to support more personalized treatment plans, reduce post-surgical complications and secondary procedures (i.e., re-excision or reconstruction), and improve overall satisfaction and quality of life [4].

Fundamentally, the healing process of the post-BCS surgical cavity follows the classic overlapping phases of reparative wound healing: hemostasis, inflammation, proliferation (or granulation), and remodeling, as demonstrated in a porcine lumpectomy model [11]. Healing begins immediately after the cavity is formed, with the surgical dead space typically filling with blood (hematoma) and/or serous fluid (seroma) [11]. A provisional fibrin matrix then forms, which lacks mechanical integrity, allowing the cavity and surrounding tissue to contract while promoting inflammation and a cytokine gradient. This gradient promotes angiogenesis, fibroblast proliferation and infiltration, and collagen deposition. As fibroblasts differentiate into myofibroblasts, they further drive tissue contraction and fibrotic remodeling, resulting in stiff scar tissue that can lead to pain, breast deformities, and altered breast consistency, all of which can negatively impact a patient’s psychological well-being [12].

Over the past three decades, mechanistic computational models have been developed to depict the wound healing process, primarily focusing on cutaneous (skin) repair. These models have evolved to better capture some of multi-scale and multiphase complexities of the healing process [13, 14]. More recently, computational models have been applied to BCS, aiming to predict post-surgical healing and cosmetic outcomes [15–17]. However, these models lack comprehensive calibration against experimental and clinical breast healing data. Furthermore, current models fail to capture the complex interactions between cellular mechanobiological activity, extracellular matrix (ECM) deposition and remodeling, and tissue contraction, all which are critical factors for accurately modeling the long-term contracture of breast tissue and overall breast deformation.

To address this gap, we previously developed and calibrated a computational mechanobiological model of breast cavity healing following BCS using porcine lumpectomy data and published clinical data [18]. This finite element model integrates biological, microstructural, and mechanical dynamics, including fibroblast infiltration, collagen remodeling, and ECM stiffening, capturing key drivers of breast tissue contraction and deformation. The model also couples nonlinear tissue mechanics with biological fields (i.e., fibroblast density and inflammation signals) described through reaction-diffusion equations. Pro-inflammatory cytokines drive fibroblast infiltration and proliferation, which in turn promote local collagen deposition, ultimately influencing tissue stiffness and active stresses contributing to contraction. Our previous simulations used a generalized breast geometry [18]. Thus, in our original work, while we were able to calibrate the dynamics of the wound healing model, we were unable to investigate the effect of patient-specific factors, namely breast shape, density, and tumor location and size.

In parallel with computational efforts, several clinical studies have evaluated how patient-specific characteristics influence BCS outcomes using statistical analyses of patient-reported data. For example, surgical decision-making models have correlated breast and tumor characteristics with post-surgical aesthetics and quality of life outcomes [19–21]. These studies show that factors such as tumor-to-breast volume ratio and tumor location can significantly affect cosmetic results and patient wellbeing, offering potential guidance on whether a patient may benefit more from BCS or mastectomy. Additional work using *BREAST* − *Q*^*TM*^ questionnaires and other surveys has revealed associations between variables such as lower breast density, higher cavity-to-breast volume percentage (CBVP), increased age, elevated body mass index (BMI), breast irradiation, and adjuvant therapy with poorer cosmetic outcomes and reduced patient satisfaction [22–27]. However, while these studies identify key trends, their reliance on questionnaire-based feedback limits insight into the underlying mechanobiological processes driving individualized healing outcomes.

In this study, we build upon our previously developed and experimentally calibrated model of post-BCS breast cavity healing [18], integrating a large dataset of preoperative magnetic resonance imaging (MRI) scans from the Cancer Imaging Archive [28] to define how individual factors, such as breast geometry, breast density, cavity volume, and cavity location, impact breast healing and deformation outcomes. Simulation results were further used to train machine learning surrogate models, enabling rapid evaluation of the effects of patient-specific features on healing dynamics and cosmetic outcomes. These patient-specific simulations and ML metamodels have the potential to support breast surgeons and patients in making more informed surgical decisions and developing personalized treatment plans, with the goal of enhancing outcome predictability, minimizing complications, and improving patient satisfaction and quality of life.

## 2 Methods and Materials

### 2.1 Preoperative MRI processing

Patient-specific data were obtained from preoperative MRI scans of breast cancer patients, sourced from The Cancer Imaging Archive, with data collected by Duke University Hospital [28]. A total of 103 patients were randomly selected from the database, excluding those with bilateral breast cancer or multiple tumors in the same breast. As displayed in Fig. 1, each MRI was processed using 3D Slicer, with tissue segmentation performed by applying intensity thresholding to estimate the volumes of adipose tissue, fibroglandular tissue, and tumor tissue [29]. Breast tissue regions were defined relative to the chest wall using MRI intensity and anatomical landmarks to distinguish boundaries from surrounding thoracic structures. Breast density for each patient was calculated as the percentage of fibroglandular tissue relative to the total breast volume. Three-dimensional breast geometries were also generated from the MRI data and meshed in 3D Slicer. Tumors were modeled as ellipsoids using dimensions provided in the repository’s datasheets. A summary of the patient and tumor characteristics is provided in Table 1.

**Fig. 1.**
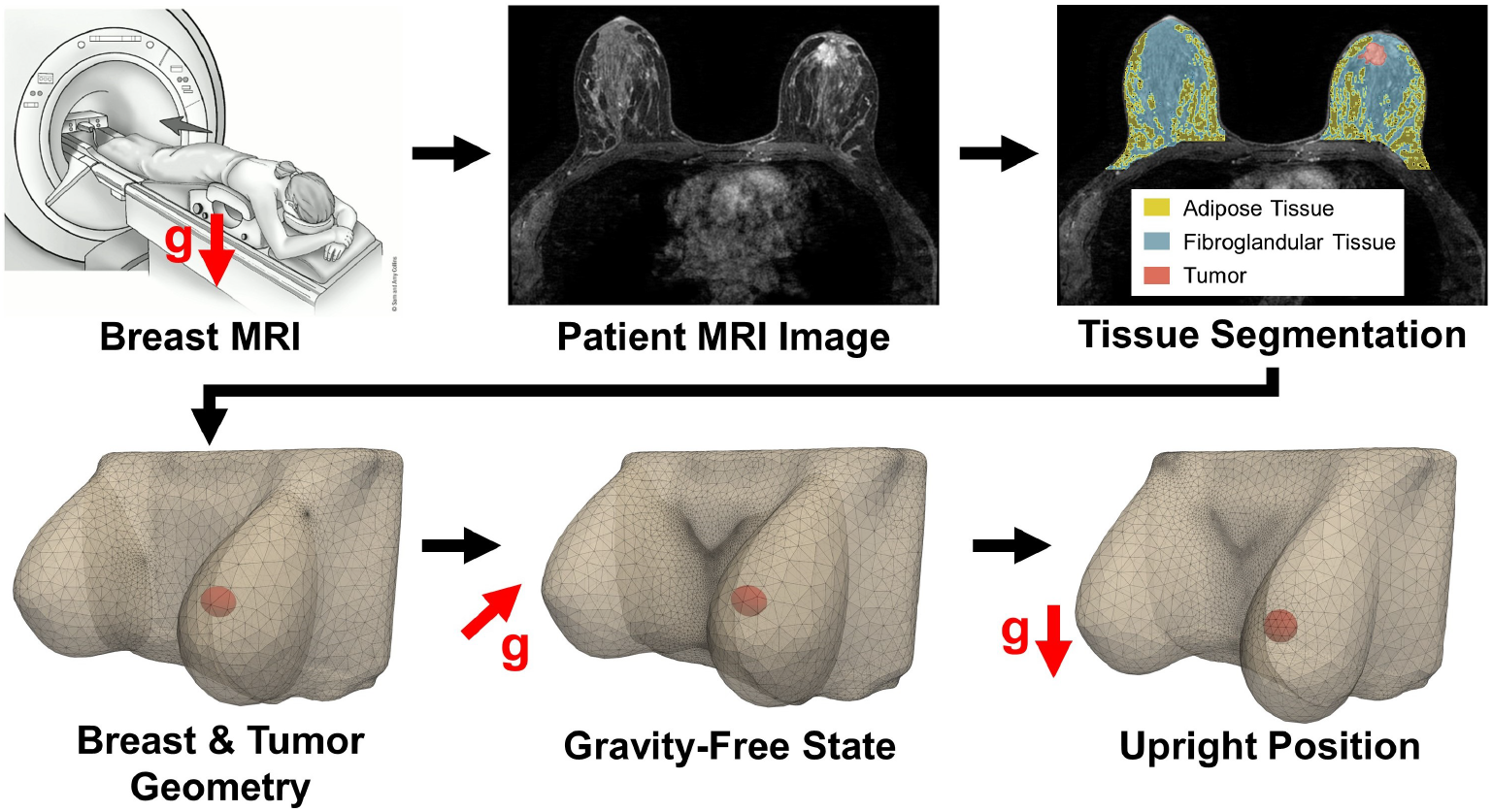
Preoperative MRI scans were processed to extract patient-specific breast and tumor characteristics. Tissue segmentation was performed on each scan to estimate the volumes of adipose tissue (yellow), fibroglandular tissue (blue), and the tumor tissue (red). These segmentations were used to construct 3D breast and tumor geometries representing the prone imaging position. Finite element simulations were then conducted to determine the unloaded (gravity-free) configuration, followed by deformation under gravity to approximate the breast and cavity geometry in the upright position. The top left panel includes an image adapted from the American Cancer Society [30].

**Table 1.** Patient breast and tumor characteristics from processed MRIs (n=103).

| Characteristic | Number of Patients (%) | Median | Range |
| --- | --- | --- | --- |
| Age (years) | - | 51.85 | 29.29–81.59 |
| Upper Outer Quadrant | 43 (41.75) | - | - |
| Upper Inner Quadrant | 30 (29.13) | - | - |
| Lower Outer Quadrant | 24 (23.30) | - | - |
| Lower Inner Quadrant | 6 (5.83) | - | - |
| Breast Density<br>(% of Fibroglandular Tissue) | - | 25.95 | 4.39–71.13 |
| Tumor Volume (mm<sup>3</sup>) | - | 1,485.32 | 33.41–35,096.57 |
| Tumor Major Axis Length (mm) | - | 19.10 | 1.95–59.93 |
| Breast Volume (mm<sup>3</sup>) | - | 755,890 | 90,191–3,208,695 |
| Tumor-to-Breast Volume (%) | - | 0.20 | 0.0015–3.86 |
| Tumor Depth* (mm) | - | 19.74 | 6.96–54.58 |

\*Tumor depth is defined as the distance from the tumor centroid to the nearest skin surface.

Since MRIs are acquired with patients lying in a prone position, the breasts are deformed under gravity, which does not reflect their natural upright shape and appearance. To correct for this, finite element simulations were performed to model the deformation of the breast and tumor as they transition from the prone to the upright position (Fig. 1). Both the breast and tumor were modeled as nearly incompressible, neo-Hookean hyperelastic materials. Material properties of the breast tissue were derived from each patient’s breast density using the rule of mixtures. Based on literature review [31] and parameter sensitivity testing, Young’s modulus values of 1.5 kPa and 6 kPa were assigned to adipose and fibroglandular tissues, respectively. The postsurgical breast cavity was modeled using the tumor geometry and location. Material properties assigned to the cavity reflected serous fluid accumulation immediately after BCS, with a shear modulus of 0 kPa and a bulk modulus of 300 kPa to simulate its near-incompressibility.

The excision volume for the 103 MRI-derived patient-specific geometries was based on standard surgical guidelines for early-stage invasive breast cancer, which recommend “no ink on tumor” and no additional negative margin width [32]. However, recognizing the variability in surgical techniques across different surgeons and patient cases, the extent of tissue removal can vary significantly. To account for this, an additional 60 geometries were generated using the same preoperative MRIs, with a randomized uniform negative cavity margin ranging from 0 mm and 5 mm, reflecting the range commonly used by surgeons [32, 33].

### 2.2 Computational model framework

The computational mechanobiological model is a custom finite element solver executed in C++, with the code repository link included at the end of the manuscript. The framework was initially developed to simulate cutaneous wound healing [34–36] and was subsequently adapted and calibrated to be specific to the breast [18]. A general overview of the model is provided below, with a more detailed description available in our previous publications [18, 35, 36]. Model parameters used in this study are consistent with those described in Harbin et al. (2023) [18], which were calibrated using both experimental porcine lumpectomy data and available human clinical data.

#### 2.2.1 Kinematics

The reference breast geometries are described with material coordinates **X** ∈ ℬ_0_ ⊂ ℝ^3^. The time-dependent configuration, ℬ_*t*_, is obtained through deformation mapping, *φ*, described as **x** = *φ*(**X**, *t*). Fibroblast density, cytokine concentration, and collagen density are defined as *ρ*(**x**, *t*), *c*(**x**, *t*), *ϕ*(**x**, *t*), respectively, with the collagen matrix described through fiber dispersion *κ*(**x**, *t*) and preferred fiber orientation **a**_0_(**x**, *t*). Local geometries changes are captured through the deformation gradient, **F** = *∂***x***/∂***X**, which can be split to capture both elastic and plastic deformation **F** = **F**^*e*^**F**^*p*^. The plastic deformation tensor can be described through an orthonormal basis [**a**_0_, **s**_0_, **n**_0_] around the preferred fiber orientation depicted as

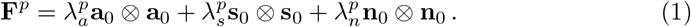

#### 2.2.2 Biochemical fields

Fibroblast density and cytokine concentration are represented as reaction-diffusion partial differential equations

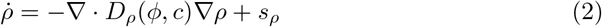

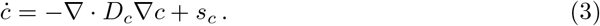

The fibroblast diffusion is dependent on both collagen density and cytokine concentration

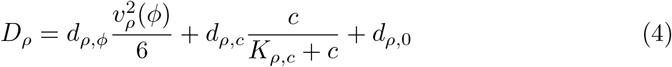

while the cytokine diffusion coefficient is assumed constant. The fibroblast and cytokine source terms are defined as

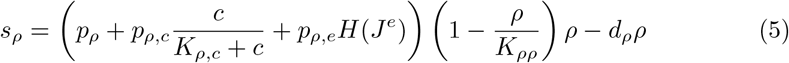

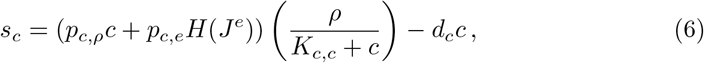

with *H*(*J*^*e*^) representing a mechanosensing activation function

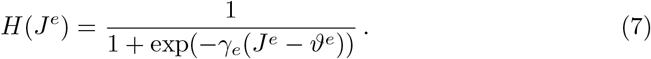

#### 2.2.3 Mechanical Equilibrium and Stress

In the absence of body forces, the balance of linear momentum is reduced to

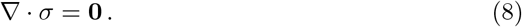

The total stress comprises both passive and active contributions

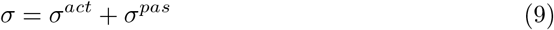

dictated by the passive material response and active extracellular matrix (ECM) contraction and reorientation directed by fibroblasts and myofibroblasts. The material behavior is described through the hyperelastic strain energy function

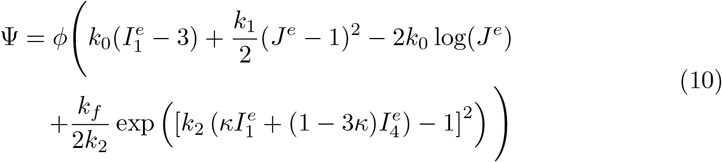

taking into account the isotropy of the non-collagenous matrix of the ECM and the anisotropy of the collagen fibers. It is parameterized by *k*_0_, *k*_1_, *k*_2_, *k*_*f*_, with the neoHookean contributions, *k*_0_ and *k*_1_, determined through each patient’s unique breast density using the rule of mixtures. Several studies were used to inform the material properties of adipose and fibroglandular tissue, as we estimate their Young’s modulus to be 10 kPa and 40 kPa, respectively [37–45].

The passive material response is also dependent on microstructure fields *ϕ, κ* and elastic invariants of the deformation 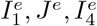. These depict the elastic volume change, *J*^*e*^ = det(**F**^*e*^), the trace of the elastic right Cauchy Green tensor, 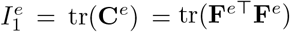, and the deformation in the preferred fiber direction, 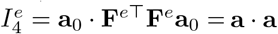, with **a** representing the deformed fiber orientation.

The passive stress is determined through the strain energy function by

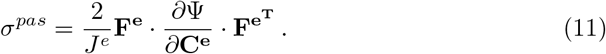

The active stress features couplings between the fibroblast density, cytokine concentration, collagen density, and preferred fiber orientation

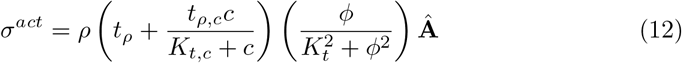

with **Â** = **A***/*tr(**A**), **A** = **I** + (1 − 3*κ*)**a** ⊗ **a**.

#### 2.2.4 Tissue Remodeling

Local remodeling of the ECM is encoded through a series of ordinary differential equations describing the microstructural fields 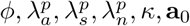. Collagen deposition is a function of both fibroblast density and cytokine concentration

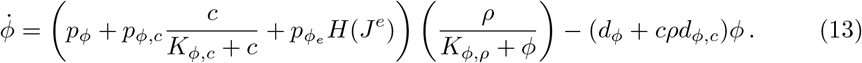

The change in plastic deformation occurs independently in the established orthonormal basis

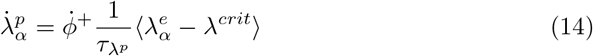

with *α* = {*a, s, n*}. It is dependent on the positive term of the collagen rate change, 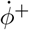, which signifies the new collagen deposition rate. The Macaulay brackets ⟨•⟩ specify that plastic deformation only occurs beyond a set threshold deformation *λ*^*crit*^.

The change in preferred collagen fiber orientation

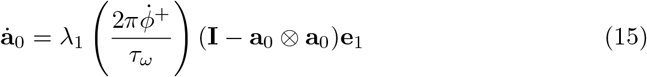

allows for the reorientation of the principal fiber direction to the direction of the maximum principal stretch, with *λ*_1_, **e**_1_ being the largest eigenvalue and corresponding eigenvector. The change in fiber dispersion

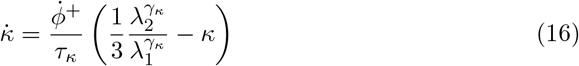

is dependent on the ratio of the first two eigenvalues, *λ*_1_ and *λ*_2_.

### 2.3 Multi-fidelity Gaussian process surrogates

#### 2.3.1 Multi-fidelity Gaussian process regression overview

The finite element model is computationally expensive, making tasks such as sensitivity analyses impractical for large-scale patient-specific simulations. Furthermore, even though the random sampling of patients from the MRI database captures the breadth of possible patient-specific characteristics, for a new patient, a full finite element simulation would still be needed. To overcome these limitations, we employed multi-fidelity Gaussian process (GP) regression to develop a computationally efficient surrogate model.

The multi-fidelity GP regression approach is adapted from Perdikaris et al. (2017) [46], and an overview is shown in Fig. 2. The framework builds on standard GP regression with an autoregressive stochastic formulation and extends it to incorporate multiple datasets from multiple fidelity levels [47–49]. It is classified as a *deep* GP, in which the posterior distribution from a lower-fidelity level is used as a training input for the higher-fidelity level. This hierarchical structure allows nonlinear correlations between fidelities to be captured effectively. By leveraging these correlations, the model combines low- and high-fidelity simulation data to enhance prediction accuracy while significantly reducing computational cost.

**Fig. 2.**
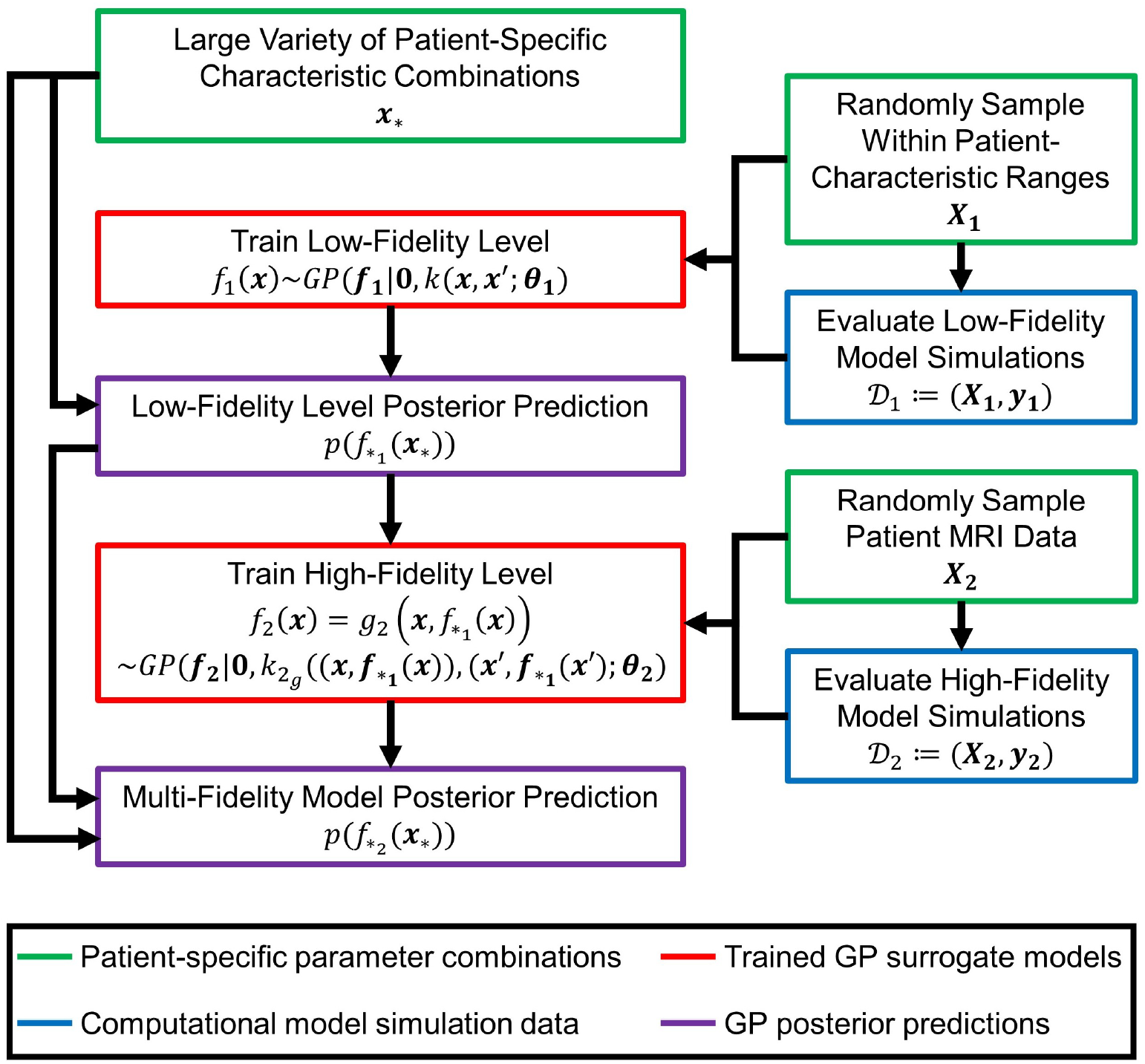
A multi-fidelity Gaussian process surrogate model was developed to evaluate how patientspecific characteristics influence BCS healing and cosmetic outcomes. The low-fidelity model consisted of 273 simulations using a generalized breast geometry with broad sampling across the patient characteristic space, while the high-fidelity model included 163 simulations based on patient-specific geometries derived from preoperative MRI scans. By leveraging correlations between the two fidelity levels, the surrogate model improves prediction accuracy despite a limited number of high-fidelity simulations. Predictions are generated by sampling the low-fidelity level posterior and using it as input for the high-fidelity surrogate to estimate outcome metrics across the patient characteristic space.

The approach assumes that there are multiple datasets of varying fidelity expressed as 𝒟_*t*_ := (**X**_**t**_, **y**_**t**_), with t denoting increasing fidelity levels (t = 1, 2). **X**_**t**_ ⊂ ℝ^*M×d*^ represents the set of *M* total inputs (**x**_**M**_), with **y**_**t**_ the corresponding outputs. To improve numerical stability, small Gaussian random noise with zero mean and variance 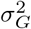 is added to the output [50]. The lowest fidelity level can be modeled using standard GP regression, with the function *f*_1_(**x**) ∼ *GP* (**f**_**1**_ | **0**, *k*(**x, x**^*′*^; *θ*_**1**_)) assigned to a zero mean GP prior and *k* being a squared exponential covariance function assigned with hyperparameters *θ*_**1**_. Hyperparameters are fit to the model by maximizing the marginal log-likelihood expressed as

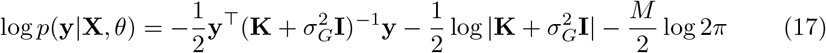

where **K** is a *M* × *M* symmetric, positive-definite matrix *K*_*ij*_ = *k*(**x**_**i**_, **x**_**j**_; *θ*). Through Bayes’ rule, the posterior distribution can be found,

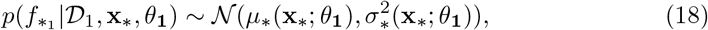

and used for new predictions for output 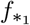 given the input **x**_*\**_. The predictive mean and variance are

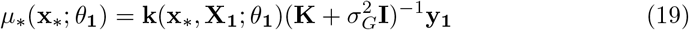

and

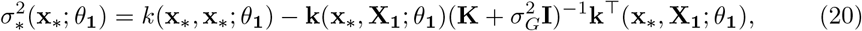

respectively, with **k**(**x**_*\**_, **X**; *θ*_**1**_) = *k*(**x**_*\**_, **x**_**1**_; *θ*_**1**_), …, *k*(**x**_*\**_, **x**_**M**_; *θ*_**1**_).

For the subsequent fidelity level, an autoregressive formulation is utilized that adds an extra dimension to the training input for the next fidelity level in the form of the GP posterior from the previous fidelity level

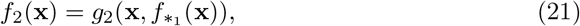

where 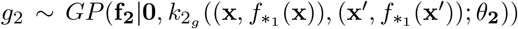. Such an approach decouples the problem into standard GP regression schemes at each fidelity level, thus capturing nonlinear correlations between fidelity levels without the limitation of computational cost. Similarly, the covariance function is also a function of the posterior of the previous fidelity level

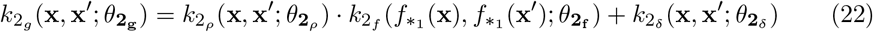

with 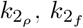, and 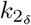 being squared exponential covariance functions along with their associated hyperparameters 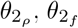, and 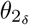. The hyperparameters can again be learned through the maximization of the marginal log-likelihood, displayed in Eq. 17. As stated above, the predictive posterior distribution for the lowest fidelity level is Gaussian with a corresponding mean and variance (Eqs. 19 and 20). The sequential fidelity level also yields a Gaussian posterior, as it is dependent on the Gaussian prediction from the first level, and is given by

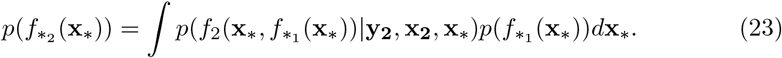

Monte Carlo integration is used to calculate the posterior predictive mean and variance for all fidelity levels following the lowest fidelity level [46].

#### 2.3.2 Multi-fidelity Gaussian process surrogate training

In addition to the 163 high-fidelity simulations based on patient-specific breast geometries, 273 low-fidelity simulations were performed using a simplified generalized breast geometry with a coarser mesh. In the low-fidelity simulations, the breast and cavity geometries were approximated as a hemisphere and a sphere, respectively, as shown in the example in Fig. 3. Four patient-specific characteristics were used as input parameters for the surrogate models: breast density, cavity volume, breast volume, and cavity depth (defined as the shortest distance between the breast surface to the center of the cavity) (Fig. 3). These features were selected for their relevance to breast tissue mechanical properties, breast geometry, and cavity geometry and location. Input values were generated using Latin hypercube sampling [51] to ensure broad coverage of the characteristic space representative of the MRI-derived patient population (Table 1).

**Fig. 3.**
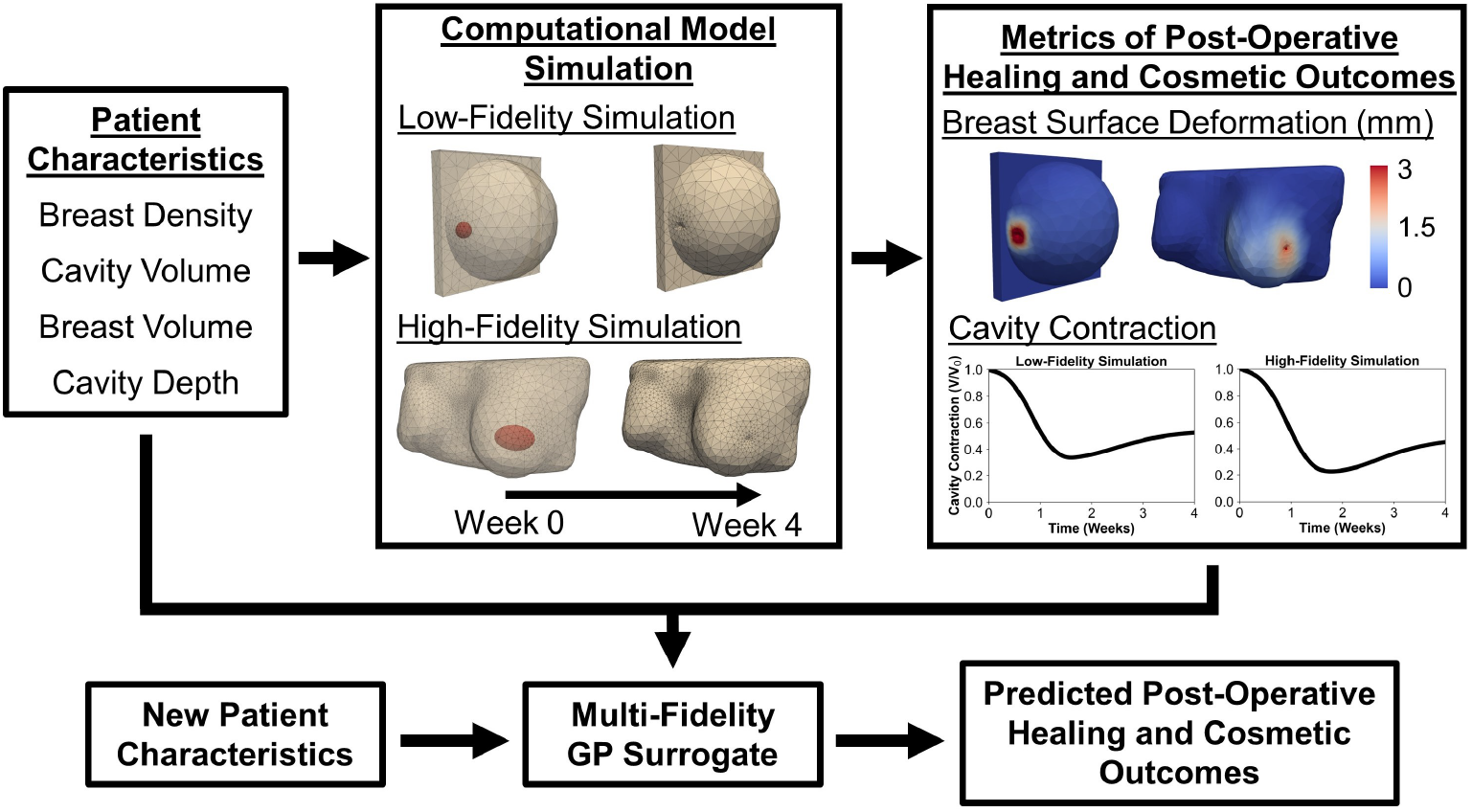
Overview of the multi-fidelity GP surrogate modeling approach used to evaluate the impact of patient-specific characteristics on post-BCS healing and cosmetic outcomes. Breast geometries were defined by four parameters: breast density, cavity volume, breast volume, and cavity depth. Finite element simulations were performed using both generalized (low-fidelity) and patient-specific (highfidelity) geometries to sample this characteristic space. Simulation outputs, including breast surface deformation and cavity contraction, were paired with input parameters to train two separate multifidelity GP surrogate models. Once trained, the surrogates could rapidly predict outcome metrics with quantified uncertainty, providing a computationally efficient alternative to full-scale simulations.

Two multi-fidelity GP surrogates were constructed as shown in Fig. 3, each trained to predict a distinct postoperative healing or cosmetic outcome metric derived from the finite element simulations (see Fig. 1). The first surrogate predicted breast surface deformation, quantified as the percentage of the cancer-affected breast surface exhibiting deformation greater than 1 mm at four weeks post-surgery (%BSD). The second surrogate predicted breast cavity contraction over the same 4-week period.

For validation, the high-fidelity dataset was randomly split into a training set (n = 150) and a testing set (n = 13). After training the surrogate models on the training set, predictions were generated for the testing set and compared to ground-truth simulation results. This procedure was repeated 20 times with different random splits, resulting in 260 unique test simulations evaluated against the surrogate predictions.

### 2.4 Qualitative assessment of post-surgical cosmetic outcomes

In addition to quantitative metrics, post-surgical cosmetic outcomes were qualitatively evaluated to assess the presence of visually noticeable breast surface deformations (i.e., divot or indentation). Five reviewers independently assessed the 163 high-fidelity patient simulations to determine whether a visible surface defect was present four weeks after surgery. Each reviewer, in a blinded fashion, compared preoperative breast surface renderings with the simulated postoperative outcomes and recorded a binary response indicating whether visible deformation was observed.

To translate the qualitative assessments into a predictive framework, reviewer responses for each patient were converted into a numerical consensus score. These scores were then used to train a GP surrogate model to predict the likelihood of visible post-surgical deformation using the same four patient-specific input features described earlier. Validation followed the same repeated-split methodology used for the multi-fidelity surrogates.

## 3 Results

### 3.1 Patient-specific model simulations capture cavity healing and breast deformation outcomes following BCS

Building upon our previous work using a generalized breast geometry to simulate post-BCS healing [18], we applied the computational model to perform 273 low-fidelity simulations (generalized geometry) and 163 high-fidelity simulations based on patient-specific breast geometries. These simulations incorporated clinically relevant variations in breast density, breast volume, cavity volume, and cavity depth. Figure 4 presents detailed finite element results for one representative patient, illustrating the multi-scale, spatiotemporal cavity healing response simulated over 4 weeks following BCS. The simulation aligns with trends observed in both our prior computational work and experimental porcine lumpectomy studies [11, 18]. Immediately following surgery, fibroblast and collagen densities at the cavity center were minimal, consistent with seroma or hematoma formation, while cytokine concentrations within the cavity peaked and subsequently declined over the four-week period [52, 53]. Fibroblast density increased sharply after week 1 and plateaued by week 4 (Fig. 4), coinciding with progressive deposition and remodeling of collagen by fibroblasts and myofibroblasts.

**Fig. 4.**
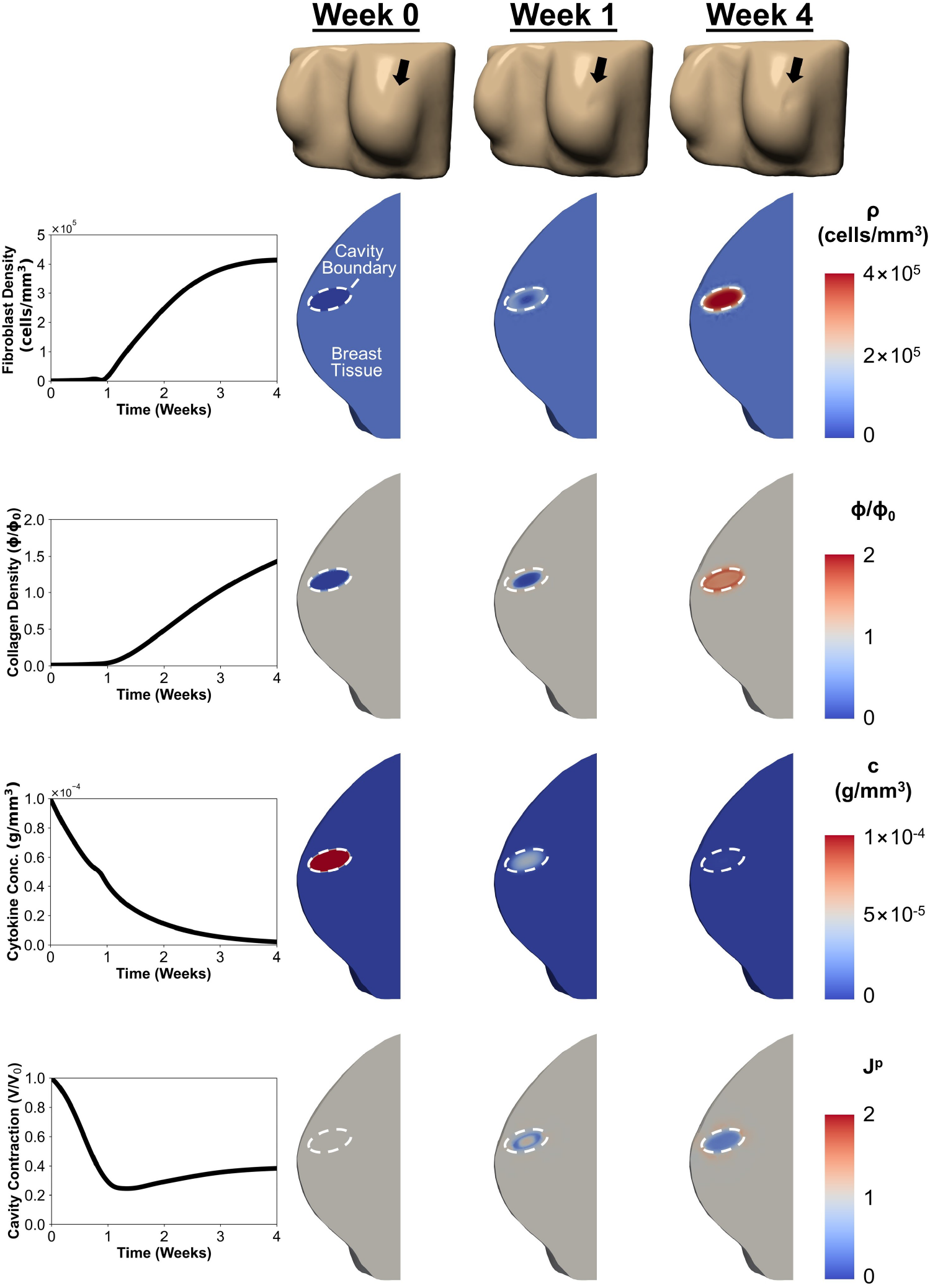
Computational simulation results for a representative patient with the following characteristics: breast volume = 644,448 mm^3^, breast density = 26.35%, cavity volume = 4,555.57 mm^3^, and cavity depth = 16.27 mm. Plots show time-dependent changes in fibroblast density, collagen density, and cytokine concentration at the cavity center, and cavity contraction. Corresponding contour plots at weeks 0, 1, and 4 illustrate healing progression. The location of breast surface deformation is indicated by a black arrow at the top of each panel.

Cavity contraction was most pronounced during the first postoperative week, reaching a minimum volume shortly thereafter, before slightly rebounding and stabilizing by week 4 (Fig. 4), a pattern consistent with both porcine and clinical data [11, 54]. Permanent deformation (*J*^*p*^) developed across the cavity region and adjacent healthy tissue, resulting in visible breast surface deformation (Fig. 4). At the time of tumor excision (t = 0 week), there was no change in tissue volume across the breast domain (*J*^*p*^ = 1). Shortly after surgery, permanent contracture (*J*^*p*^ *<* 1) developed at the tissue-cavity interface, causing moderate breast surface deformation by week 1 (Fig. 4). By week 4, extensive permanent contracture was evident throughout the cavity domain, while surrounding tissue experiences tensional forces (*J*^*p*^ *>* 1) oriented perpendicular to the cavity surface (Fig. 4). This differential deformation resulted in pronounced breast surface deformation adjacent to the cavity (Fig. 4). These findings are consistent with established tissue repair and scar formation mechanisms, where fibroblasts and myofibroblasts within the cavity contract and reorient newly deposited collagen fibers while exerting tensile forces on the surrounding ECM [36, 55].

A comparison of patient-specific simulation outcomes for breast surface deformation and cavity contraction, shown in Fig. 5, highlights the impact of individual patient characteristics, including breast density (Fig. 5A), cavity volume (Fig. 5B), breast volume (Fig. 5C), and cavity depth (Fig. 5D). Patients with lower breast density exhibited more pronounced surface deformation, including a more pronounced surface indentation, compared to those with higher breast density (Fig. 5A). Lower breast density was also associated with more rapid and severe cavity contraction (Fig. 5A). Larger cavities led to significantly greater surface deformation (Fig. 5B). While larger cavities experienced greater overall contraction, smaller cavities contracted more rapidly during the early healing phase (Fig. 5B). When evaluating the effect of breast volume, patients with larger breast volumes experienced greater magnitudes of surface deformation, however, the %BSD was similar across breast sizes (Fig. 5C). Cavity contraction followed similar trends in both cases, though patients with smaller breast volumes demonstrated slightly greater contraction by week 4 (Fig. 5C). The impact of cavity depth was also evident, with more superficial cavities leading to greater surface deformation compared to deeper cavities (Fig. 5D). Although both cases exhibited similar cavity contraction trends, patients with more superficial cavities showed marginally greater contraction by week 4 (Fig. 5D).

**Fig. 5.**
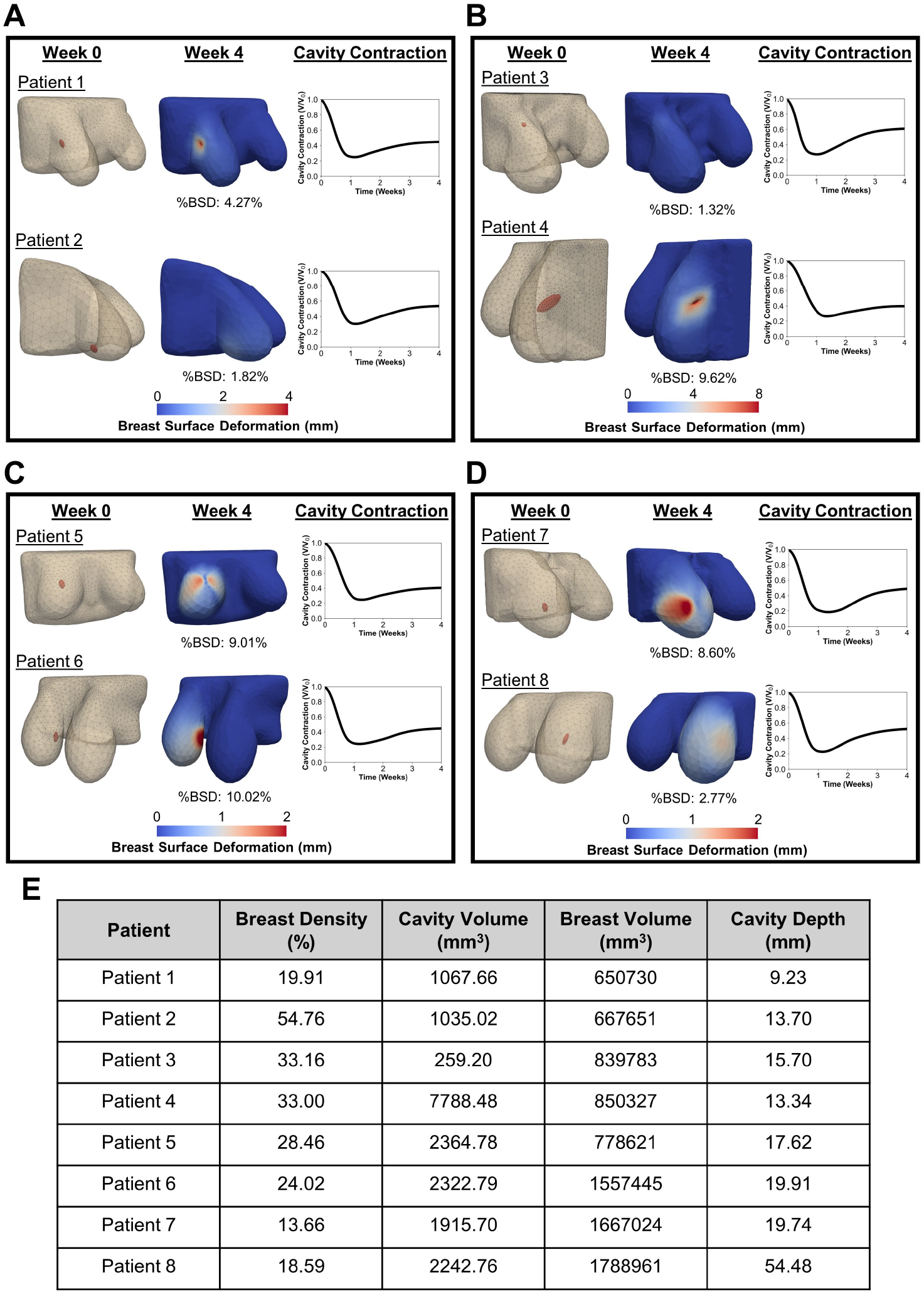
Simulation results illustrating breast surface deformation and cavity contraction for patients with differing **A** breast density, **B** cavity volume, **C** breast volume, and **D** cavity depth, while other patient characteristics are similar. The corresponding values for each patient are summarized in panel **E**. For each case, contours show the initial breast and cavity geometry, the breast surface deformation 4 weeks post-surgery, and the time course of cavity contraction. The %BSD metric represents the percentage of breast surface area that experienced deformation more than 1 mm.

Breast surface deformation and cavity contraction data were extracted from each patient-specific simulation and used to train the GP surrogate models. A summary of the 163 patient-specific simulation outcomes is shown in Fig. 6, highlighting key metrics related to postoperative healing and cosmetic outcomes following BCS. On average, breast cavity volume decreased rapidly during the first 9 days post-surgery, contracting to approximately 25.70% of its original volume (Fig. 6A). The shaded 95% credible interval during this period ranged from 18.61% to 34.83% (Fig. 6A), indicating moderate variability across patients. Following this initial contraction phase, cavity volume steadily increased and stabilized, with a broader credible interval reflecting greater interpatient variability (Fig. 6A). By week 4, the average cavity volume was approximately 51.64% of the originally excised volume, with the distribution of cavity contraction values at week 4 shown in Fig. 6B. For breast surface deformation, a significant portion of patients experienced minimal to no deformities, with 55 patients predicted to have less than 1% of the breast surface deform by more than 1 mm (Fig. 6C). These predictions were consistent with the qualitative assessment, where 88 patients were unanimously judged to have no visible post-surgical deformation (Fig. 6D). In contrast, 41 patient simulations showed more than 10% of the breast surface deforming over 1 mm, indicating more pronounced contour changes and poorer cosmetic outcomes (Fig. 6C). These findings aligned with the qualitative assessment, where 38 of these patients were unanimously judged to exhibit visible surface deformation (Fig. 6D).

**Fig. 6.**
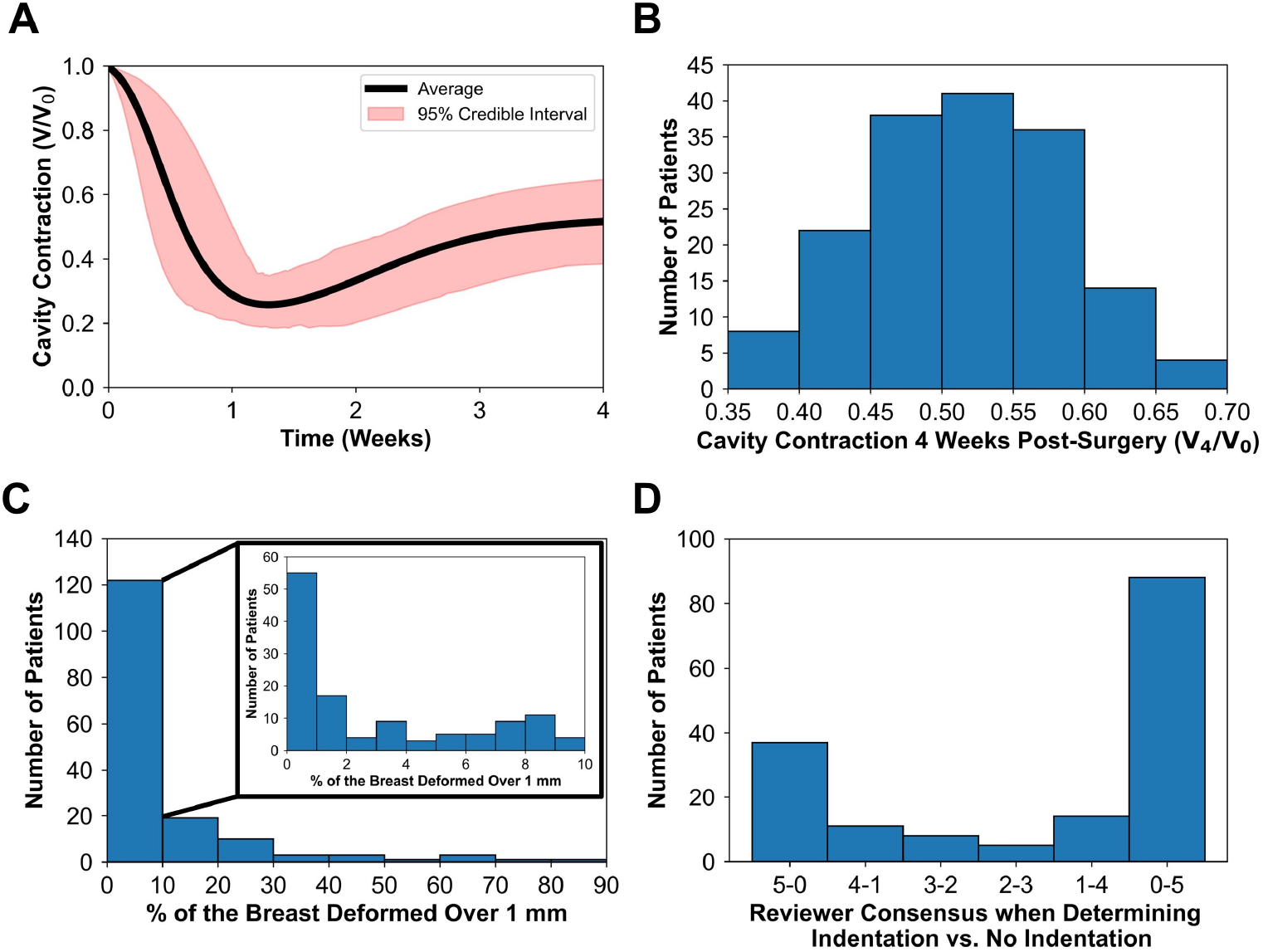
Summary of predictive postoperative cavity healing and breast surface deformation outcomes from all patient-specific simulations. **A** Average cavity contraction over time, with shaded areas representing the 95% credible interval. **B** Cavity contraction at 4 weeks post-BCS for each patient. **C** Percentage of the breast surface area deformed by more than 1 mm at 4 weeks post-BCS (%BSD). **D** Reviewer consensus from qualitative assessment, indicating whether a visible deformation (i.e., divot or indentation) was present on the breast surface at 4 weeks post-surgery.

Using the simulation data, we calibrated and validated multi-fidelity GP surrogate models. For each of 20 iterations, the high-fidelity dataset was randomly divided into 150 training and 13 testing samples. Surrogate predictions for the 260 testing samples were compared to computational model outputs. Validation showed that the %BSD surrogate achieved a mean absolute error (MAE) of approximately 3.48%, while the cavity contraction surrogate had a root mean square error (RMSE) of 0.046. Additionally, when quantitative surrogate predictions of breast surface deformation were compared to the reviewer assessment consensus, the model showed agreement in approximately 81.23% of cases.

### 3.2 Patient characteristics influence post-surgical breast surface deformation

After calibrating the GP surrogate models, they were used to predict how variations in patient-specific breast and cavity characteristics affect postoperative healing and cosmetic outcomes. Specifically, the four patient characteristics, breast volume, breast density, cavity volume, and cavity depth, were varied to determine their impact on breast surface deformation. By sampling patient characteristic values within the feasibility ranges reported in Table 1, 2,500 predictive simulations were performed. These were used to evaluate two outcomes: %BSD (Fig. 7(i)), and the likelihood of visible post-surgical deformation (Fig. 7(ii)), both as functions of the four patient characteristics.

**Fig. 7.**
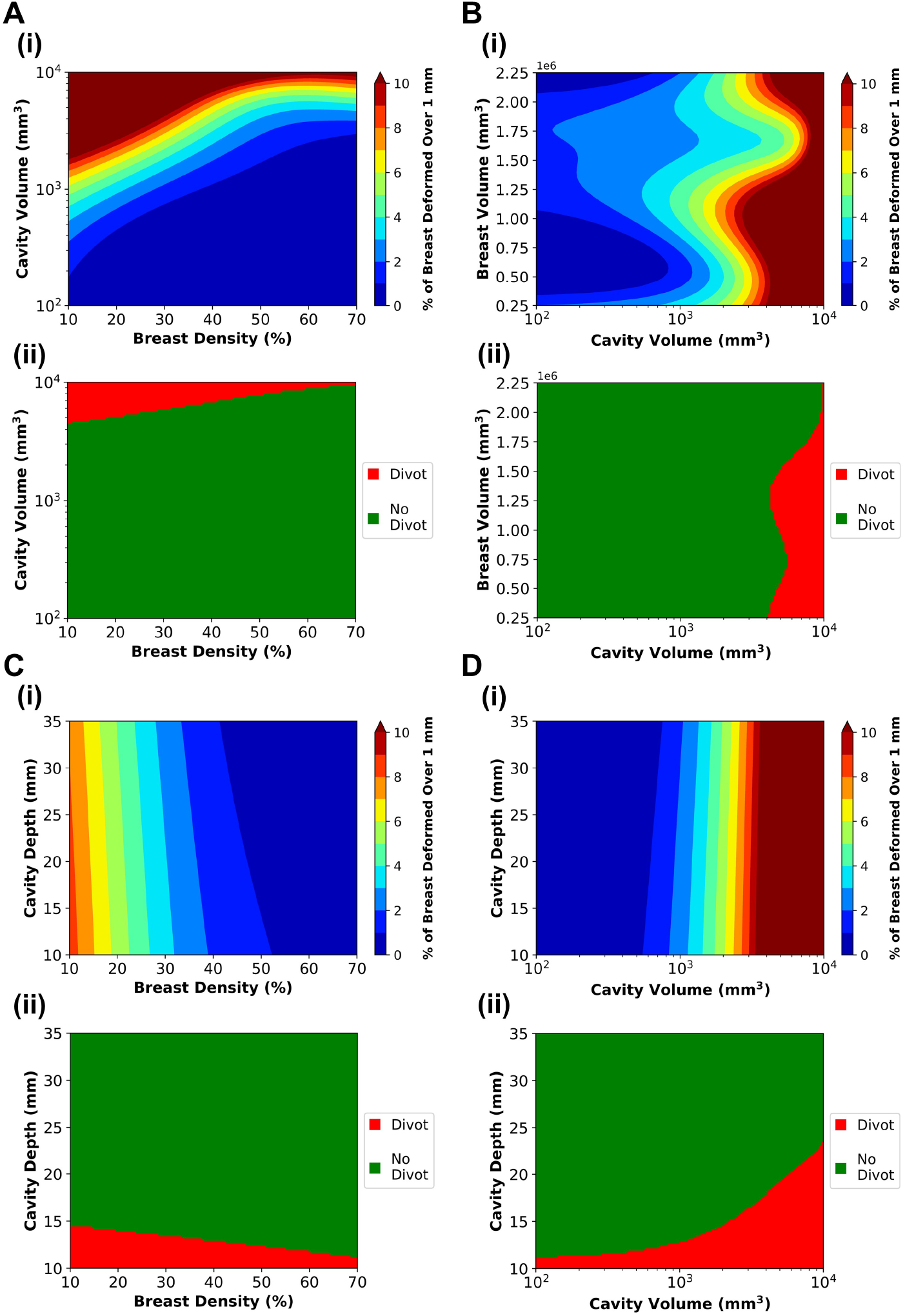
Relationship between breast density, cavity volume, breast volume, and cavity depth and their influence on breast surface deformation. Two outcome metrics are shown: **(i)** the percentage of the breast surface deforming more than 1 mm (%BSD), and **(ii)** the presence of visible post-surgical deformation (i.e., divot or indentation). Plots were generated using predictions from the multi-fidelity Gaussian process surrogate model and the Gaussian process classifier, respectively, by varying two of the four patient characteristics at a time. Median values for the remaining characteristics, derived from MRI data and summarized in Table 1, were held constant during the analysis.

Consistent with clinical reports [23–26, 56–58], both breast density and cavity volume significantly impacted breast surface deformation. Lower breast densities (i.e., breasts consisting of primarily fatty tissue or scattered regions of fibroglandular tissue) and larger cavity volumes were associated with greater deformation risk (Fig. 7A(i) and B(i)). The contour plot in Fig. 7A(i) indicates that for patients with low breast density, cavity volume and breast density have comparable influence on surface deformation. In contrast, for patients with higher fibroglandular tissue (*>* 50%), deformation is more strongly influenced by cavity volume. As shown in (Fig. 7B(i)), breast volume had a lesser effect, likely due to its correlation with breast density and excision volume [26, 57–59]. Surface deformation was more pronounced for superficial cavities (i.e., those with smaller depth values), as shown in (Fig. 7C(i) and D(i)), consistent with clinical observations [60]. However, overall cavity depth had a lesser influence on surface deformation compared to breast density and cavity volume.

Trends observed in the qualitative predictions of visible surface deformations (Fig. 7(ii)) largely mirrored those from the quantitative analyses, with some notable variations in how patient characteristics influenced deformation risk. The contour plots of cavity volume versus breast density (Fig. 7A(ii)) and breast volume versus cavity volume (Fig. 7B(ii)) showed similar patterns to their quantitative counterparts (Fig. 7A(i),B(i)), indicating that larger cavity volumes and lower breast densities were associated with a higher likelihood of visibly identifiable surface deformations. While the quantitative analysis suggested that breast density had a greater impact than cavity depth on surface deformation (Fig. 7C(i)), the qualitative predictions indicated a more balanced influence between the two (Fig. 7C(ii)). Specifically, lower values of both breast density and cavity depth increased the likelihood of an observable deformation. Similarly, although cavity volume was previously identified as the dominant factor in influencing breast surface deformation (Fig. 7D(i)), the qualitative analysis revealed that for smaller cavity volumes, cavity depth had a greater influence, while cavity volume became more impactful as its size increased (Fig. 7D(ii)).

### 3.3 Patient characteristics influence the time-dependent contraction of the breast cavity

Using the same approach applied to breast surface deformation, the cavity contraction multi-fidelity surrogate model was utilized to predict how variations in patient-specific characteristics influenced cavity contraction over time. For each characteristic, 2,500 predictive simulations were generated by sampling across the previously established parameter ranges. These results are summarized in the contour plots in Fig. 8, illustrating the time-dependent effects of patient factors on cavity contraction.

**Fig. 8.**
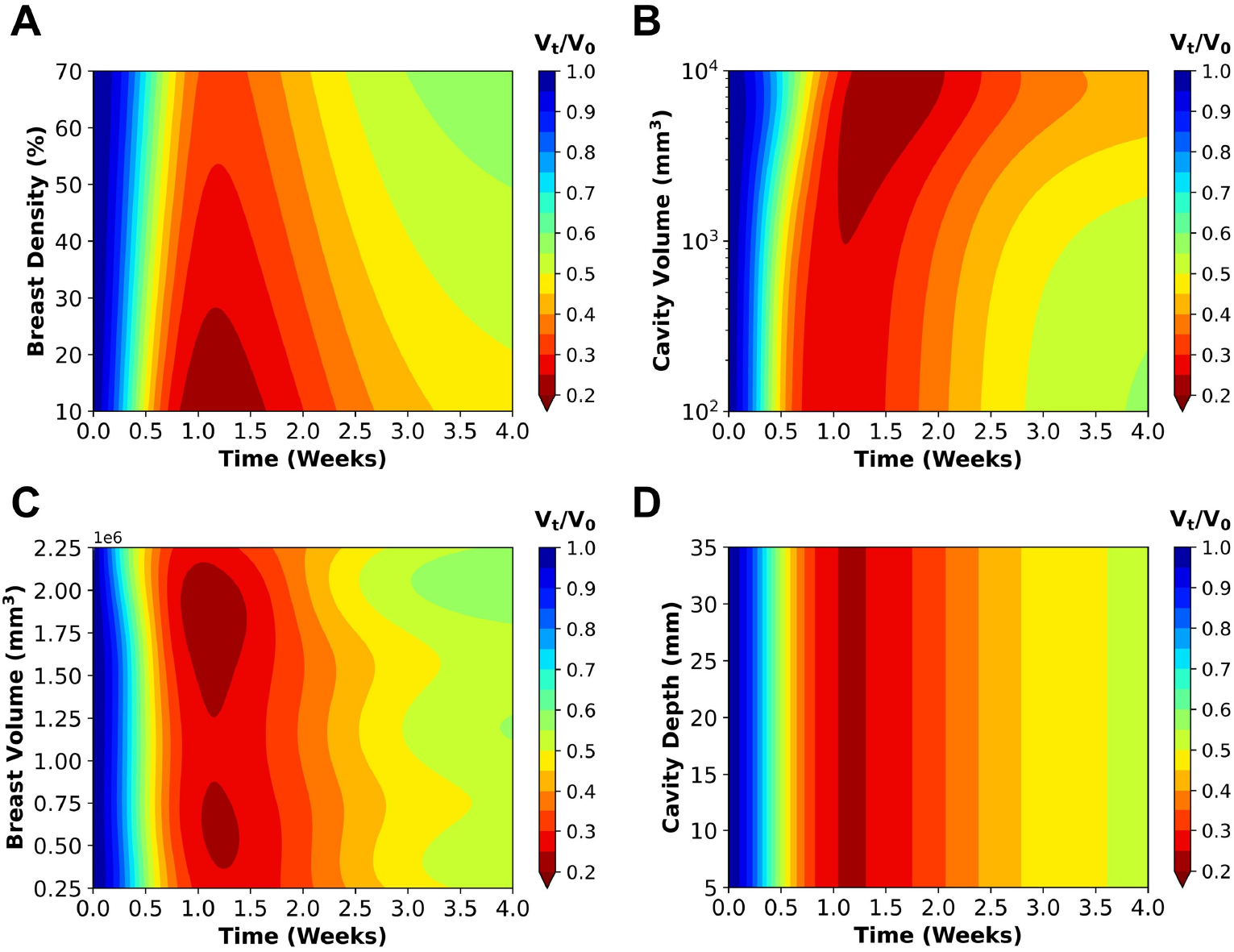
Time-dependent effects of **A** breast density, **B** cavity volume, **C** breast volume, and **D** cavity depth on predicted cavity contraction. Plots were generated using the multi-fidelity GP surrogate model by varying one parameter at a time to isolate its influence. The remaining patient characteristics were held constant at their median values, derived from processed MRI data and summarized in Table 1.

Breast cavities in low breast density regions contracted more rapidly during the first postoperative week and exhibited greater overall contraction severity over time compared to cavities in higher density (fibroglandular-rich) breasts (Fig. 8A). Similarly, larger cavity volumes were associated with greater cavity volume reduction, though smaller cavities contracted at faster rates (Fig. 8B), consistent with reported trends in human wound contraction [61, 62]. As with surface deformation, the influence of breast volume on cavity contraction was variable (Fig. 8C), likely due to interactions with breast density and excised tissue percentage [26, 57–59]. In contrast, cavity depth had little effect on either the rate or extent of cavity contraction over time (Fig. 8D).

## 4 Discussion

BCS provides the advantage of preserving breast tissue and aesthetics, but its outcomes can vary greatly due to the complexities of wound healing and individual patient factors. Notably, BCS is associated with a significant incidence of post-surgical breast deformities, with approximately one-third of women developing indentations or breast asymmetry [20, 23, 26, 63]. Poor cosmetic outcomes can have a lasting impact on a patient’s self-esteem and quality of life, potentially affecting emotional well-being and overall recovery [4]. Variability in surgical outcomes and their impact on patient wellbeing have driven interest in personalized medicine, which guides decision-making for breast surgeons by tailoring treatment options to individual patient characteristics. For instance, risk calculators and decision models applied in clinical practice can provide valuable insights to guide treatment choices based on prior patient data. One example is a risk calculator developed by Jonczyk et al. (2021), which estimates a patient’s unique risk of acute complications following BCS, oncoplastic surgery, and various mastectomy procedures, taking into account patient demographics and other risk factors [64]. Similarly, Vos et al. developed an informed decision model that estimates the likelihood of achieving a good cosmetic outcome after BCS compared to mastectomy based on the patient’s tumor-to-breast volume ratio and tumor location [19, 20]. Additionally, Pukancsik et al. (2017) created a surgical decision model for

BCS, oncoplastic surgery, and mastectomy, focusing on the location and extent of tissue excised [21]. Although these decision models are based on statistical correlations, they still do not fully capture the wound healing process or how individual patient characteristics specifically influence healing outcomes. To address this gap, we incorporated individual patient MRIs into a predictive breast healing mechanobiological model that was previously calibrated against experimental and clinical data [18]. By running simulations with the calibrated model over a large patient database from the Cancer Imaging Archive [28], we sought to advance the mechanistic understanding of how patient-specific factors impact outcomes following BCS.

The computational model used captures multi-scale cell and tissue mechanics, including complex couplings between mechanobiological activity, collagen deposition and remodeling, and deformation and biomechanics of the whole breast [18, 34–36]. To date, there are a limited number of models that depict the healing of deep soft tissue wounds. For example, Garbey and collaborators developed a 2D model based on a single patient geometry using cytokine signaling concepts from Javierre et al. (2009), adapted to predict time-dependent breast shape changes after BCS but without accounting for permanent tissue remodeling [15, 16, 65]. Vavourakis et al. (2016) developed a 3D finite element model, integrating soft tissue biomechanics with mathematical descriptions of inflammation and angiogenesis introduced by Sherratt and Murray (1991) [17, 66]. Their model was calibrated with MRI and optical surface scans from four patients taken before and 6 to 12 months after surgery [17, 66]. In contrast, our framework was calibrated against porcine and clinical data with an emphasis on (myo)fibroblast activity and collagen remodeling. These couplings enable the explanation of macroscale changes in tissue mechanics and elastoplastic deformation over long periods of time. Even though we do not model a detailed inflammatory cascade or angiogenesis, we do incorporate basic inflammation dynamics through our cytokine concentration field, similar to prior cutaneous wound healing models [14, 65, 67–69].

A key contribution of this work is the application of the calibrated 3D model to over 100 patient-specific geometries derived from preoperative MRI scans available in the Cancer Imaging Archive [28], allowing for the assessment of how patient-specific characteristics influence healing and cosmetic outcomes using a multi-fidelity GP surrogate modeling approach. These MRI scans were used to simulate individualized wound healing outcomes in 163 3D finite element simulations. The patient-specific simulations were computationally expensive, often requiring several days to complete for each individual. Thus, it was difficult to generalize from patient-specific geometries to general population insights. To enhance the dataset, low-fidelity simulations were conducted using a generalized breast geometry and a coarser mesh. This ultimately allowed the use of multi-fidelity GP regression to make efficient predictions. This approach capitalizes on the correlations between low- and high-fidelity data, out-performing models trained exclusively on high-fidelity data [46]. This methodology has previously been successfully employed by our group for the modeling and prediction of tissue expansion outcomes, demonstrating its efficacy in capturing complex mechanobiological processes with reduced computational effort [70, 71].

A similar modeling and surrogate-based approach was used by Zolfagharnasab et al. (2018), who simulated breast deformation following BCS using the model proposed by Vavourakis et al. (2016) across a limited set of patient characteristics [17, 72]. However, their study was constrained by a small sample size of patient MRIs, broad assumptions about patient-specific factors, a simplified wound healing model, and limited analysis regarding the influence of patient-specific characteristics [72]. In contrast, our multi-fidelity surrogate model was trained on over 400 finite element simulations that span a wide range of anatomical and surgical variations, more accurately reflecting the heterogeneity seen in the Cancer Imaging Archive MRI dataset [28]. While our earlier work relied on a generalized breast geometry derived from population averages [18], this study advances that foundation by incorporating patient-specific geometries from preoperative MRIs. This allowed for a more detailed investigation of how individual patient features affect healing trajectories and deformation outcomes. Our analysis revealed that breast density and cavity volume were more influential than breast volume in determining breast surface deformation (Fig. 7B), with cavity volume exerting a stronger effect in patients with denser breasts, and both parameters contributing equally in those with fattier breasts (Fig. 7A). These results are consistent with prior clinical studies [23–26, 56–60], while also expanding on existing knowledge by revealing how multiple patient-specific features interact to influence post-surgical outcomes.

For our analysis, breast surface deformation was evaluated using two complementary metrics: a quantitative measure, represented through %BSD, and a qualitative assessment, evaluating the presence of visible post-surgical surface deformation. This dual approach addressed the inherently subjective nature of cosmetic outcome assessments in clinical settings, aligning computational model predictions with both clinician and patient perspectives. The qualitative assessment offered a more intuitive and clinically relevant interpretation, with the presence of a visible divot or indentation serving as a direct and tangible indicator of surface deformation, complementing the quantitative data. Among the key findings of this comparative analysis, the quantitative measure revealed that breast density and cavity volume had a greater influence on breast surface deformation than cavity depth (Fig. 7C(i) and D(i)). In contrast, the qualitative assessment showed that cavity depth was more influential than breast density in predicting visible surface deformations (Fig. 7C(ii)). Additionally, cavity depth had a greater impact than cavity volume for smaller cavities, while their influence on surface deformation was similar for larger cavities (Fig. 7D(ii)). These findings highlight the value of integrating both quantitative and qualitative metrics to provide a more comprehensive understanding of breast surface deformation, bridging the gap between computational analysis and clinical evaluation.

This study is not without limitations. While the computational model simulations incorporated patient-specific data from preoperative MRIs to characterize breast and tumor features, longitudinal or postoperative imaging was not available to corroborate model predictive simulations against individual healing outcomes. Although the model was calibrated using data from our previous work [18], mechanobiological parameters likely vary between individuals and would benefit from future calibration using longitudinal, patient-specific data. Our analysis focused on breast surface deformation and cavity contraction as indicators of healing and cosmetic outcomes. Expanding this framework to include additional clinically relevant metrics, such as nipple-areolar complex asymmetry, will likely provide a more comprehensive assessment of postoperative aesthetics. The current model also does not account for radiation therapy, which is commonly administered in the weeks following BCS [73, 74], and has been associated with fibrosis and poorer cosmetic outcomes [26, 27, 75, 76]. Future extensions of the model could incorporate additional biological processes, including angiogenesis and immune cell-cytokine interactions, further enhancing its predictive power. Ultimately, this framework may support the development and in silico testing of therapeutic approaches such as regenerative breast tissue fillers.

In conclusion, this work presents a computational mechanobiological model of cavity healing following BCS that incorporates patient-specific data to predict individual healing outcomes and evaluate the influence of key patient characteristics on postsurgical healing and cosmetic results. By integrating preoperative MRIs with finite element simulations, we assessed how variations in breast density, cavity volume, breast volume, and cavity depth affect healing dynamics and postoperative deformation. Simulation results were used to train single- and multi-fidelity GP surrogate models, enabling efficient prediction of cavity contraction and breast surface deformation across a diverse patient population. The framework advances current approaches by not only forecasting individualized healing trajectories four weeks following surgery, but also capturing tissue remodeling and deformation, offering a mechanistic understanding of post-BCS outcomes. The model has the potential to inform personalized treatment planning, helping surgeons and patients anticipate healing responses, reduce the risk of poor cosmetic outcomes, and ultimately enhance quality of life for breast cancer survivors.

## Data Availability

All data is available at https://github.com/zharbin/2025_BCS

## Supplementary information

The finite element model is available in the following repository: https://github.com/zharbin/2025_BCS.

## Acknowledgements

This work was supported by an NSF CMMI Multiscale Mechanobiology of Growth and Remodeling During Wound Healing grant (A.B.T.; 1911346). Z.H. was a recipient of the Leslie Bottorff Fellowship.

## Declarations

- Disclosures: The authors (Z.H., S.L.H, A.B.T) declare that a provisional patent application has been filed related to the computational model and code described in this work.

## Notes

### Competing Interest Statement

The authors (ZJH, SLH, ABT) declare that a provisional patent application has been filed related to the computational model and code described in this work.

### Author Declarations

This study used only openly available de-identified human patient data from the Cancer Imaging Archive https://wiki.cancerimagingarchive.net/pages/viewpage.action?pageId=70226903

