## Supplemental Table 1 for "Computational modeling of patient-specific healing and deformation outcomes following breast-conserving surgery based on MRI data"

**Table 1 Parameters for the computational mechanobiological model.**

| Parameter | Description | Value | Reference |
|---|---|---|---|
| $\rho_0$ $[cells/mm^3]$ | Nominal Fibroblast Density | 55051 | [1] |
| $d_{\rho,\phi}$ $[-]$ | Fibroblast Diffusion Scaling Constant | 1582.3 | [1] |
| $\Delta$ $[-]$ | Skewness of Fibroblast Speed $v_\rho(\phi)$ | 0 | [1] |
| $d_{\rho,c}$ $[mm^2/hr]$ | Cytokine-Increased Fibroblast Diffusivity | $6.12 \times 10^{-3}$ | [1] |
| $d_{\rho,0}$ $[mm^2/hr]$ | Baseline Fibroblast Diffusivity | $6.12 \times 10^{-5}$ | [1, 2] |
| $p_\rho$ $[1/hr]$ | Fibroblast Proliferation | $9 \times 10^{-4}$ | [1] |
| $p_{\rho,c}$ $[1/hr]$ | Cytokine-Increased Proliferation | 0.015314 | [1] |
| $K_{\rho,c}$ $[-]$ | Proliferation Saturation due to Cytokine | $1 \times 10^{-5}$ | [3] |
| $p_{\rho,e}$ $[1/hr]$ | Mechanoregulation of Fibroblast Proliferation | $p_\rho/2$ | [4] |
| $K_{\rho,\rho}$ $[-]$ | Fibroblast Division Saturation | $10 * \rho_0$ | [5] |
| $d_\rho$ $[1/hr]$ | Fibroblast Death Rate | $p_\rho(1-\rho_0/K_{\rho\rho})$ | [5] |
| $c_0$ $[g/mm^3]$ | Initial Cytokine Concentration Inside Cavity | $1 \times 10^{-4}$ | [3] |
| $D_c$ $[mm^2/hr]$ | Cytokine Diffusion Coefficient | 0.01208 | [6–8] |
| $p_{c,\rho}$ $[1/hr]$ | Fibroblast Secretion of Cytokine | $1.635 \times 10^{-18}$ | [3] |
| $p_{c,e}$ $[1/hr]$ | Mechanoregulation of Cytokine | $5.45 \times 10^{-18}$ | [3] |
| $K_{c,c}$ $[mol/mm^3]$ | Cytokine Saturation | 1 | [3] |
| $d_c$ $[1/hr]$ | Cytokine Death Rate | 0.005 | [1] |
| $k_0$ $[MPa]$ | Linear Stiffness | $[3.356 \times 10^{-3}, 1.342 \times 10^{-2}]$ | Dependent on Patient-Specific Breast Density |
| $k_1$ $[MPa]$ | Compressibility | $[0.1667, 0.6667]$ | Dependent on Patient-Specific Breast Density |
| $k_f$ $[MPa]$ | Fiber Stiffness | 0.015 | [9] |
| $k_2$ $[-]$ | Nonlinear Stiffening | 0.048 | [9] |
| $\gamma_e$ $[-]$ | Shape of Mechanosensing Curve | 5 | [3] |
| $\vartheta_e$ $[-]$ | Midpoint of Mechanosensing Curve | 2 | [3, 10] |
| $t_\rho$ $[MPa]$ | Contractile Force of Fibroblasts | $2.33548 \times 10^{-7}$ | [1] |
| $t_{\rho,c}$ $[MPa]$ | Contractile Force of Myofibroblasts | $3.28571 \times t_\rho$ | [1] |
| $K_{t,c}$ $[-]$ | Traction Saturation due to Cytokine | $1 \times 10^{-5}$ | [3] |
| $K_t$ $[-]$ | Saturation of Mechanical Force by Collagen | 0.2 | [1] |
| $p_\phi$ $[1/hr]$ | Collagen Production | $1.4 \times 10^{-8}$ | [1] |
| $p_{\phi,c}$ $[1/hr]$ | Collagen Production Activated by Cytokine | $7.0 \times 10^{-8}$ | [1] |
| $K_{\phi,c}$ $[-]$ | Collagen Production Saturation due to Cytokine | $1 \times 10^{-4}$ | [3] |
| $p_{\phi_e}$ $[1/hr]$ | Collagen Production Activated by Stretch | $p_\phi$ | [3] |
| $K_{\phi,\rho}$ $[-]$ | Collagen Production Saturation due to Collagen Fraction | 1.06 | [3] |
| $d_\phi$ $[1/hr]$ | Collagen Degradation | $p_\phi(\rho_0/(K_{\phi,\rho}+1))$ | [10] |
| $d_{\phi,c}$ $[1/hr]$ | Collagen Degradation Activated by Cytokine | $8.81 \times 10^{-5}$ | [11] |
| $\tau_{\lambda^p}$ $[1/hr]$ | Rate of Plastic Deformation | 0.05 | [1] |
| $\tau_\omega$ $[hr]$ | Time Constant for Reorientation | $10/(K_{\phi,\rho}+1)$ | [3] |
| $\tau_\kappa$ $[hr]$ | Time Constant for Dispersion | $1/(K_{\phi,\rho}+1)$ | [3] |
| $\gamma_\kappa$ [-] | Shape of Dispersion Rate Curve | 2 | [3] |
